# Associations between prenatal caffeine exposure and child development: Longitudinal results from the Adolescent Brain Cognitive Development (ABCD) Study

**DOI:** 10.1101/2024.06.18.24309117

**Authors:** Hailey Modi, David AA Baranger, Sarah E Paul, Aaron J Gorelik, Alana Hornstein, Jared V Balbona, Arpana Agrawal, Janine D Bijsterbosch, Ryan Bogdan

## Abstract

**Objective:** Though caffeine use during pregnancy is common, its longitudinal associations with child behavioral and physical health outcomes remain poorly understood. Here, we estimated associations between prenatal caffeine exposure, body mass index (BMI), and behavior as children enter adolescence.

**Method:** Longitudinal data and caregiver-reported prenatal caffeine exposure were obtained from the ongoing Adolescent Brain and Cognitive Development (ABCD)**^SM^** Study, which recruited 11,875 children aged 9-11 years at baseline from 21 sites across the United States starting June 1, 2016. Prenatal caffeine exposure was analyzed as a 4-level categorical variable, and further group contrasts were used to characterize “any exposure” and “daily exposure” groups. Outcomes included psychopathology characteristics in children, sleep problems, and BMI. Potentially confounding covariates included familial (e.g., income, familial psychopathology), pregnancy (e.g., prenatal substance exposure), and child (e.g., caffeine use) variables.

**Results:** Among 10,873 children (5,686 boys [52.3%]; mean [SD] age, 9.9 [0.6] years) with nonmissing prenatal caffeine exposure data, 6,560 (60%) were exposed to caffeine prenatally. Relative to no exposure, daily caffeine exposure was associated with higher child BMI (β=0.08; FDR-corrected *p*=0.02), but was not associated with child behavior. Those exposed to two or more cups of caffeine daily (n=1,028) had greater sleep problems than those with lower/no exposure (β>0.92; FDR-corrected *p*<0.04).

**Conclusion:** Daily prenatal caffeine exposure is associated with heightened childhood BMI, and when used multiple times a day greater sleep problems even after accounting for potential confounds. Whether this relationship is a consequence of prenatal caffeine exposure or its correlated factors remains unknown.

## Introduction

As the most widely used psychoactive substance in the world, caffeine is commonly consumed during pregnancy, with more than 45% of pregnant Americans reporting daily use^1^. Although evidence generally supports the safety of moderate caffeine consumption in adults^2^, the safety of use during pregnancy remains poorly understood. As caffeine has a 3-4 fold increased half-life among pregnant people (i.e., 11-23 hours)^3^, readily crosses the placenta^4^, and is metabolized more slowly by the fetus^5^, higher accumulations of caffeine may adversely impact pregnancy and child health and development. Indeed, meta-analyses suggest that caffeine consumption during pregnancy, especially during later stages, is associated with miscarriage, stillbirth, and other adverse birth outcomes (e.g., low birth weight, premature birth, small for gestational age)^6,7^, even when under the 200 mg dose (i.e., two cups of moderate-strength coffee) considered safe by the American College of Obstetricians and Gynecologists^8^. Other studies linking prenatal caffeine exposure to shorter height^9^ and heavier weight^10^ during early childhood raise concern that effects may extend into later life.

Accumulating, but equivocal, evidence has linked prenatal caffeine exposure to behavioral problems during childhood. Non-human animal studies have found that moderate-high caffeine administration (i.e., the equivalent of 2-3 cups/day) during pregnancy slows neural migration, decreases neuronal numbers, and induces cognitive deficits among offspring^11^. A human study in a cohort of 1,083 mother-child pairs found that children of mothers who consumed more than 200 mg of caffeine per day during pregnancy performed worse on IQ tests at 5.5 years of age^12^. However, a separate study in a cohort of 2,000 found no difference in behavioral measures in children who were exposed to caffeine prenatally as compared to controls^13^. More recently, Zhang and colleagues found that prenatal caffeine exposure was associated with externalizing behavior, but not internalizing problems or cognition, during middle childhood in the Adolescent Brain and Cognitive Development (ABCD) Study^14^. However, the persistence and severity of these differences as children enter adolescence, as well as the potential emergence of other related differences, has remained unexplored.

Here, using longitudinal data from the ABCD Study^®^, we test whether prenatal caffeine exposure is associated with behavioral problems (i.e., internalizing, externalizing, attention, thought, and social problems, as well as psychotic-like experiences [PLEs], sleep problems) and body mass index (BMI) as children enter adolescence. We hypothesized that any caffeine exposure would be unassociated with these phenotypes, but that daily and higher exposure would be associated with greater psychopathology, sleep problems, and BMI. We further expected that these associations would become stronger as children progress through adolescence, when greater variability in these phenotypes typically manifest.

## Methods

### Participants

The Adolescent Brain and Cognitive Development (ABCD)^SM^ Study is a longitudinal study of complex behavior and biology from middle childhood to late adolescence/young adulthood. The ABCD Study^®^ recruited 11,875 children (ages 8.9-11; born between 2005-2009) at baseline (June 1, 2016 - October 15, 2018) from 21 research sites across the United States (https://abcdstudy.org/sites/abcd-sites.html). The study includes a family-based design in which twins (n = 2,108), triplets (n = 30), non-twin siblings (n = 1,589), and singletons (n = 8,148) were recruited. We used data (release 5.0; https://abcdstudy.org/) from the baseline, 1-year follow-up (1YFU; phenotypic data; n_1-year follow-up recruited_ = 11,199; Dates: 8/30/17-3/1/20), 2-year follow-up (2YFU; phenotypic; n_2-year follow-up recruited_ = 10,066; Dates: 7/30/18-9/27/21), and 3-year follow-up (3YFU; phenotypic data; n_3-year follow-up recruited_ = 9,508; Dates: 8/7/19-1/15/22) sessions^15^. All parents or caregivers (10,131 of 11,875 biological mothers [85.3%]) provided written informed consent, and children provided verbal assent to a research protocol approved by the institutional review board at each data collection site. After accounting for missing data, analytic Ns ranged from 9,970-10,873 (**Table 1**).

**Table 1:** ABCD Study Sample Characteristics.

| Variable | Daily prenatal caffeine exposure (n = 2773) | Weekly prenatal caffeine exposure (n = 2112) | Monthly prenatal caffeine exposure (n = 1675) | No prenatal caffeine exposure (n = 4313) | Total (N = 10873) |
| --- | --- | --- | --- | --- | --- |
| Child variables |  |  |  |  |  |
| Age, mean (SD), y | 9.9 (0.6) | 9.9 (0.6) | 9.9 (0.6) | 9.9 (0.6) | 9.9 (0.6) |
| Girls | 1307 (47.2) | 1029 (48.8) | 800 (47.8) | 2051 (47.6) | 5187 (47.7) |
| Race/ethnicity<sup>b</sup> |  |  |  |  |  |
| White | 2240 (80.8) | 1772 (83.9) | 1315 (78.5) | 2893 (67.1) | 8220 (75.6) |
| Black | 480 (17.3) | 350 (16.6) | 279 (16.7) | 1098 (25.5) | 2207 (20.3) |
| Native American | 113 (4.1) | 84 (4.0) | 56 (3.3) | 119 (2.8) | 372 (3.4) |
| Pacific Islander | 11 (0.4) | 12 (0.6) | 15 (0.9) | 29 (0.7) | 67 (0.6) |
| Asian | 137 (4.9) | 109 (5.2) | 111 (6.6) | 305 (7.1) | 662 (6.1) |
| Hispanic (n = 10737) | 491 (17.7) | 329 (15.6) | 326 (19.5) | 1042 (24.2) | 2188 (20.1) |
| Other | 156 (5.6) | 92 (4.4) | 112 (6.7) | 354 (8.2) | 714 (6.6) |
| Child lifetime substance exposure |  |  |  |  |  |
| Has tried alcohol (n = 10470) | 712 (25.7) | 535 (25.3) | 384 (22.9) | 837 (19.4) | 2468 (23.5) |
| Has tried tobacco (n = 10020) | 27 (1.0) | 15 (0.7) | 8 (0.5) | 16 (0.0) | 66 (0.7) |
| Caffeine use score, mean (SD) (n = 10261) | 2.46 (6.08) | 2.15 (6.97) | 2.03 (6.75) | 1.69 (5.74) | 2.03 (6.26) |
| Pregnancy and family variables |  |  |  |  |  |
| Unplanned pregnancy (n = 10796) | 1181 (42.6) | 767 (36.3) | 596 (35.6) | 1546 (35.8) | 4090 (37.9) |
| Maternal age at birth, mean (SD), y (n = 10760) | 29.7 (6.2) | 29.7 (6.0) | 29.5 (5.7) | 29.3 (6.4) | 29.5 (6.2) |
| Prenatal vitamin use (n = 10721) | 2564 (92.5) | 2021 (95.7) | 1613 (96.3) | 4056 (94.0) | 10254 (95.6) |
| Week learned pregnancy, mean (SD) (n = 9970) | 7.3 (7.1) | 6.4 (5.5) | 6.7 (6.5) | 6.9 (7.0) | 6.9 (6.7) |
| Maternal education, mean (SD), y (n = 10861) | 15.1 (2.6) | 15.6 (2.3) | 15.5 (2.4) | 15.1 (2.7) | 15.3 (2.5) |
| Household income (n = 9980), \$ | | | | | |
| ≤ 49 999 | 812 (29.3) | 456 (21.6) | 372 (22.2) | 1274 (29.5) | 2914 (29.2) |
| 50 000 – 74 999 | 357 (12.9) | 295 (14.0) | 232 (13.9) | 501 (11.6) | 1385 (13.9) |
| 75 000 – 99 999 | 372 (13.4) | 302 (14.3) | 240 (14.3) | 525 (12.2) | 1439 (14.4) |
| 100 000 – 199 999 | 731 (26.4) | 669 (31.7) | 545 (32.5) | 1124 (26.1) | 3069 (30.8) |
| ≥ 200 000 | 289 (10.4) | 253 (12.0) | 179 (10.7) | 452 (10.5) | 1173 (11.6) |
| Family history of psychopathology<sup>c</sup> (n = 10869) |  |  |  |  |  |
| Psychosis | 81 (2.9) | 36 (1.7) | 36 (2.2) | 93 (2.2) | 246 (2.3) |
| Depression | 1008 (36.4) | 721 (34.1) | 503 (30.0) | 1109 (25.7) | 3341 (30.7) |
| Anxiety | 398 (14.4) | 256 (12.1) | 200 (11.9) | 449 (10.4) | 1303 (12.0) |
| Antisocial behavior | 489 (17.6) | 276 (13.1) | 167 (10.0) | 420 (9.7) | 1352 (12.4) |
| Mania | 190 (6.9) | 101 (4.8) | 83 (5.0) | 196 (4.5) | 570 (5.2) |
| Prenatal substance exposure before knowing of pregnancy |  |  |  |  |  |
| Alcohol (n = 10486) | 851 (30.7) | 608 (28.8) | 484 (28.9) | 755 (17.5) | 2698 (25.7) |
| Tobacco (n = 10820) | 677 (24.4) | 292 (13.8) | 170 (10.1) | 326 (7.6) | 1465 (13.5) |
| Cannabis (n = 10772) | 231 (8.3) | 118 (5.6) | 90 (5.4) | 167 (3.9) | 606 (5.6) |
| Prenatal substance exposure after knowing of pregnancy |  |  |  |  |  |
| Alcohol (n = 10821) | 135 (4.9) | 69 (3.3) | 23 (1.4) | 47 (1.1) | 274 (2.5) |
| Tobacco (n = 10840) | 340 (12.3) | 91 (4.3) | 49 (2.9) | 74 (1.7) | 554 (5.1) |
| Cannabis (n = 10833) | 101 (3.6) | 23 (1.1) | 27 (1.6) | 43 (1.0) | 209 (1.9) |
| Child primary outcomes of interest, mean (SD) |  |  |  |  |  |
| Psychotic-like experiences | 2.53 (3.5) | 2.43 (3.3) | 2.82 (3.7) | 2.59 (3.6) | 2.61 (3.6) |
| Internalizing symptoms per CBCL | 5.29 (6.0) | 5.05 (5.4) | 4.95 (5.3) | 4.89 (5.3) | 5.02 (5.5) |
| Externalizing symptoms per CBCL | 4.84 (6.3) | 4.37 (5.7) | 4.29 (5.6) | 4.20 (5.6) | 4.41 (5.8) |
| Attention symptoms per CBCL | 3.11 (3.6) | 2.98 (3.5) | 2.94 (3.4) | 2.86 (3.4) | 2.96 (3.5) |
| Thought symptoms per CBCL | 1.74 (2.4) | 1.61 (2.1) | 1.61 (2.1) | 1.54 (2.1) | 1.61 (2.2) |
| Social symptoms per CBCL | 1.68 (2.4) | 1.62 (2.2) | 1.52 (2.1) | 1.59 (2.2) | 1.61 (2.3) |
| Sleep problems per SDSC | 37.43 (8.7) | 36.80 (8.2) | 36.58 (7.6) | 35.72 (8.1) | 36.50 (8.2) |
| BMI (n = 10843) | 19.08 (4.3) | 18.74 (4.1) | 18.57 (4.0) | 18.8 (4.3) | 18.82 (4.2) |

Abbreviations: ABCD, Adolescent Brain and Cognitive Development; BMI, body mass index (calculated as weight in kilograms divided by height in meters squared); CBCL, Child Behavior Checklist; SDSC, Parent Sleep Disturbance Scale for Children.
<sup>a</sup>Data included in analyses were required to have a response of daily, weekly, monthly, or “none” for maternal report of using caffeine during pregnancy. All variables reflect measures assessed during the baseline session when children were a mean (SD) age of 9.9 (0.6) years. The estimates reported above refer to the non-winsorized versions of each variable (refer to Supp. 1 for descriptive information of the winsorized data).
<sup>b</sup>Race/ethnicity variables were coded as non-mutually exclusive dichotomous variables; as such, these numbers do not sum to 100% as participants could be included in multiple categories.
<sup>c</sup>Family history of depression, psychosis, mania, antisocial behavior, and anxiety among first-degree relatives. Psychotic-like experiences were measured by the Prodromal Questionnaire-Brief Child Version total score.

## Measures

### Prenatal Caffeine Exposure

Parents/caregivers retrospectively reported on maternal use of caffeine (No, less than once/week, more than once/week but not daily, once a day or more) during pregnancy according to the following question: “Did you/biological mother have any caffeine during pregnancy (from conception until delivery)?” Follow-up questions inquired about how much (i.e., number of cups containing caffeine) caffeine was used during pregnancy per day/week/month. As there was a large amount of missingness for this quantity assessment among those reporting caffeine use during pregnancy (∼20%), prenatal caffeine exposure was coded as 4 mutually exclusive groups: 1) no (n = 4,313), 2) monthly (n = 1,675), 3) weekly (n = 2,112), and 4) daily (n = 2,773) exposure. Additional groups were coded according to: 1) any caffeine exposure (i.e., daily, weekly, or monthly; n=6,560) and 2) multiple cups/day (i.e., ≥2 cups/day) exposure (n=1,028). All analyses were rerun excluding respondents who were not the biological mother (**Supplementary Methods**).

### Outcome Measures

#### Psychopathology. CBCL

The Child Behavior Checklist^16^ was used to assess broad-spectrum internalizing and externalizing problems as well as attention (associated with ADHD), thought (associated with psychosis), and social (associated with Autism) problems in children according to parent/caregiver report at baseline, 1YFU, 2YFU, and 3YFU. Higher scores are reflective of more problems. ***PQ-BC.*** The Prodromal Questionnaire-Brief Child Version^17,18^ total score was used to assess child-reported PLEs at baseline, 1YFU, 2YFU, and 3YFU. Higher scores indicate more PLEs.

#### Sleep

The Sleep Disturbance Scale for Children^19^ total score was used to assess sleep problems in children according to parent or caregiver report at baseline, 1YFU, 2YFU, and 3YFU. Higher scores reflect more sleep problems.

#### Body Mass Index

Child BMI was calculated using measured height and weight. As there were high levels of missingness in follow-up height and weight measurements, BMI was only calculated at baseline.

All continuous outcome measures were standardized and winsorized to ±3 SDs and are described in greater detail within **Supplementary Methods** and **Table 1**.

### Covariates

Covariates included in analyses are consistent with those used in prior analyses of prenatal caffeine and other exposures in the ABCD dataset^14,20^. The following fixed-effect covariates were dummy coded: race/ethnicity (White, Black, Asian, Hispanic, Native American, Pacific Islander, and “other”), first-degree familial history of psychopathology (alcohol addiction, drug addiction, depression, psychosis, anxiety, mania, and antisocial behavior), prenatal exposure to tobacco (0/1), alcohol (0/1), and other drugs (e.g., cocaine, heroin, oxycontin; 0/1) before or after maternal knowledge of pregnancy, unplanned pregnancy, prenatal vitamin use, child caffeine use, child substance use (alcohol and tobacco), child sex, and twin or triplet status. Annual household income was treated as a 5-level categorical variable. The following continuous covariates were included: interview age, age^2^, maternal educational level, and pubertal development score. These variables were reported by caregivers and/or participants. Adult Self-Report^21^ summary scores for caregiver externalizing, internalizing, attention, and thought problems at baseline were also included as continuous covariates. All continuous covariates were standardized.

Consistent with prior work^20^, uncommon substance use among children (i.e., use other than trying alcohol or tobacco, n=409 [e.g., having a marijuana puff or a full alcoholic drink]) or by women while they were pregnant (n=102), as well as extreme premature birth (ie, <32 weeks; n=149) and non-biological mother caregiver report (n=1,617), were not included as covariates; post hoc analyses excluding individuals based on these variables were conducted and yielded conclusions consistent with the primary analyses (**Supp. 1**; **Tables S1-S8).**

### Statistical Analysis

Values on continuous predictor and outcome variables were winsorized (to ±3 SD) prior to analyses to minimize the influence of extreme values^22^. All analyses were conducted using linear mixed-effects models (lme4 R [version 4.2.1] package^23^) with random intercepts for participant, site, and family membership. Across all models, fixed effects covariates described above were included. Omnibus tests were conducted using log-likelihood ratio tests (stats R [version 4.2.1] package^24^) between models containing prenatal caffeine use as an independent 4-level categorical variable, and models without, in order to identify outcomes that showed significant differences across groups. Post hoc contrast analyses were then performed for outcomes showing a significant association with prenatal caffeine exposure in the omnibus test: **1)** daily vs. no exposure, **2)** weekly vs. no exposure, **3)** monthly vs. no exposure, **4)** daily vs. weekly, **5)** daily vs. monthly, **6)** weekly vs. monthly, with additional contrasts for **1)** any exposure (daily, weekly, or monthly; n=6,560) vs no exposure (n=4,317), **2)** daily vs. all lower levels of exposure (n=8,100), and **3)** Multiple-cup daily exposure (i.e., ≥2 cups/day; n=1,028) vs. all lower levels of exposure (n=9,467), given evidence that adverse effects may only arise at higher doses of caffeine exposure^11,12^. FDR correction was used to adjust for multiple comparisons across outcomes (8 outcomes, two tests per outcome - omnibus test and multi-cup daily exposure vs. all lower, 16 tests total), and to adjust for multiple comparisons across post-hoc tests for each outcome (8 post-hoc contrasts per outcome of interest). Additional models including age and age^2^ interactions with prenatal exposure group (i.e., prenatal exposure group x age; prenatal exposure group x age^2^) were run to examine whether there were age-related changes in associations between prenatal caffeine exposure and child behavior as children progressed through adolescence. To test for significant interactions, likelihood ratio tests were used to compare models with interactions to those without interactions (e.g., any prenatal caffeine exposure vs any prenatal caffeine exposure x age interactions). FDR correction was used to adjust for testing multiple contrasts for each phenotype (8 tests per outcome of interest). Post-hoc contrasts (8 tests for age and age squared respectively) were similarly used to identify which forms of use contributed to the interaction.

## Results

### Prenatal Caffeine Exposure

Among 10,873 children (52.3% boys; 53.4% White; mean±SD age, 9.9±0.6 years) with non-missing data, 6,560 (60.33%) were prenatally exposed to caffeine (**Table 1**). Of these, 2,773 (25.50% of the total sample) were exposed daily during pregnancy; 2,112 (19.42% of the total sample) were exposed weekly; and 1,675 (15.41% of the total sample) were exposed monthly. Of children prenatally exposed to caffeine on a daily basis (n=2,773), 2,395 (86%) had nonmissing data on the amount of daily exposure (1 cup/day n=1,367 [57%]; ≥2 cups/day n=1,028 [43%]; mean∓SD amount 1.75∓2.33 cups/day).

### Prenatal Caffeine Exposure Associations with Child Phenotypes

#### BMI

Log-likelihood ratio tests showed a significant association between prenatal caffeine exposure and baseline BMI (χ^2^=12.57; FDR-corrected *p*=0.045; **Table S1**). Post-hoc contrasts of prenatal caffeine exposure groups revealed that: 1) BMI was significantly higher in the daily prenatal caffeine exposure group relative to the no exposure group, and 2) daily prenatal exposure was also associated with higher BMI when compared to monthly use (all βs > 0.08; all FDR-corrected *p*<0.04; **Table 2**; **Figure 1**). Weekly prenatal exposure was associated with heightened BMI when compared to the no exposure group at nominal significance levels unadjusted for multiple testing (β=0.06; *p*=0.049; FDR-corrected *p*=0.13; **Table 2**; **Figure 1**).

**Table 2:**
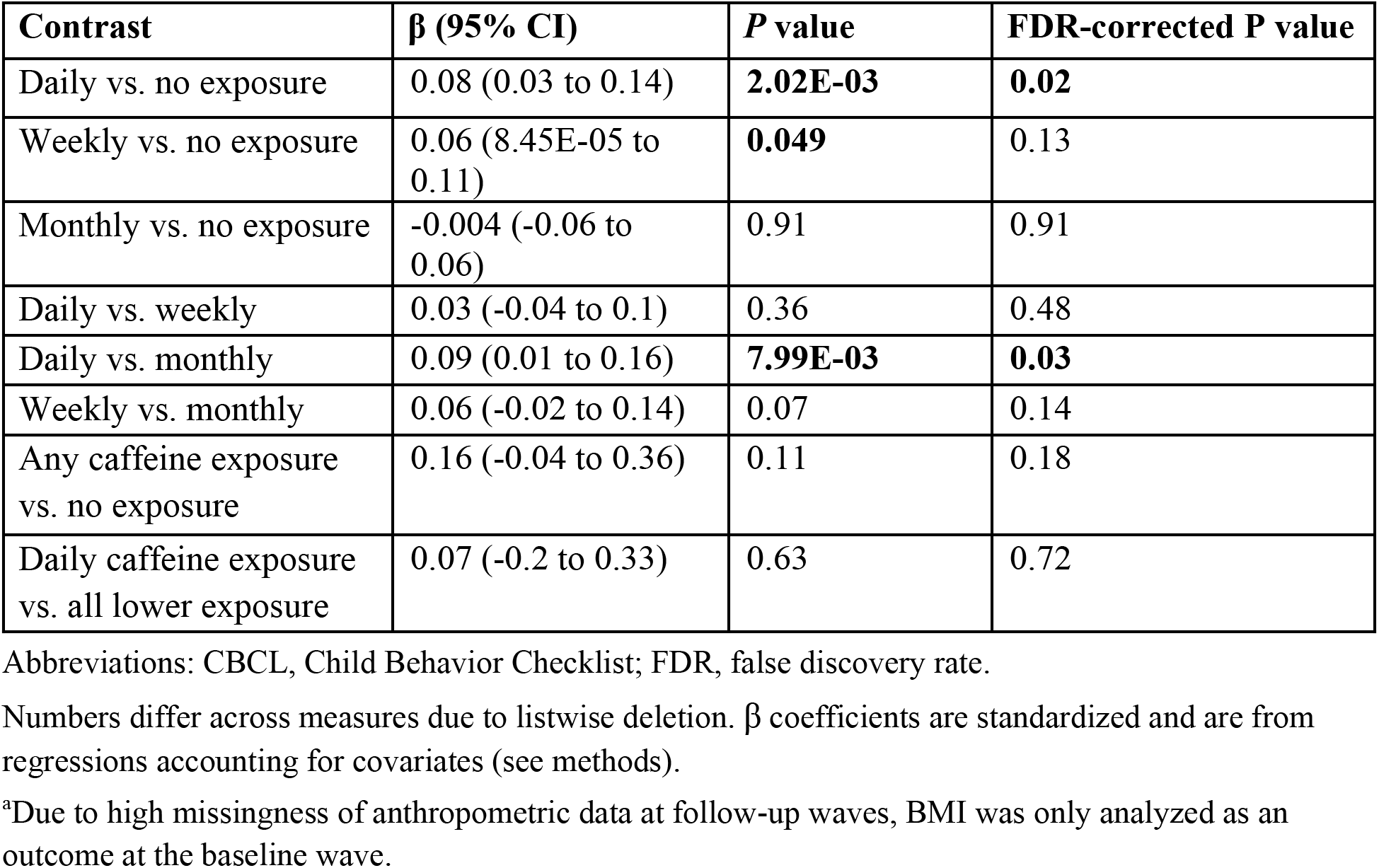
Associations Between Prenatal Caffeine Exposure and Body Mass Index^a^.

| **Contrast** | **$\beta$ (95% CI)** | ***P* value** | **FDR-corrected *P* value** |
| --- | --- | --- | --- |
| Daily vs. no exposure | 0.08 (0.03 to 0.14) | **2.02E-03** | **0.02** |
| Weekly vs. no exposure | 0.06 (8.45E-05 to 0.11) | **0.049** | 0.13 |
| Monthly vs. no exposure | -0.004 (-0.06 to 0.06) | 0.91 | 0.91 |
| Daily vs. weekly | 0.03 (-0.04 to 0.1) | 0.36 | 0.48 |
| Daily vs. monthly | 0.09 (0.01 to 0.16) | **7.99E-03** | **0.03** |
| Weekly vs. monthly | 0.06 (-0.02 to 0.14) | 0.07 | 0.14 |
| Any caffeine exposure vs. no exposure | 0.16 (-0.04 to 0.36) | 0.11 | 0.18 |
| Daily caffeine exposure vs. all lower exposure | 0.07 (-0.2 to 0.33) | 0.63 | 0.72 |

Abbreviations: CBCL, Child Behavior Checklist; FDR, false discovery rate.
Numbers differ across measures due to listwise deletion. $\beta$ coefficients are standardized and are from regressions accounting for covariates (see methods).
<sup>a</sup>Due to high missingness of anthropometric data at follow-up waves, BMI was only analyzed as an outcome at the baseline wave.

**Table 3:**
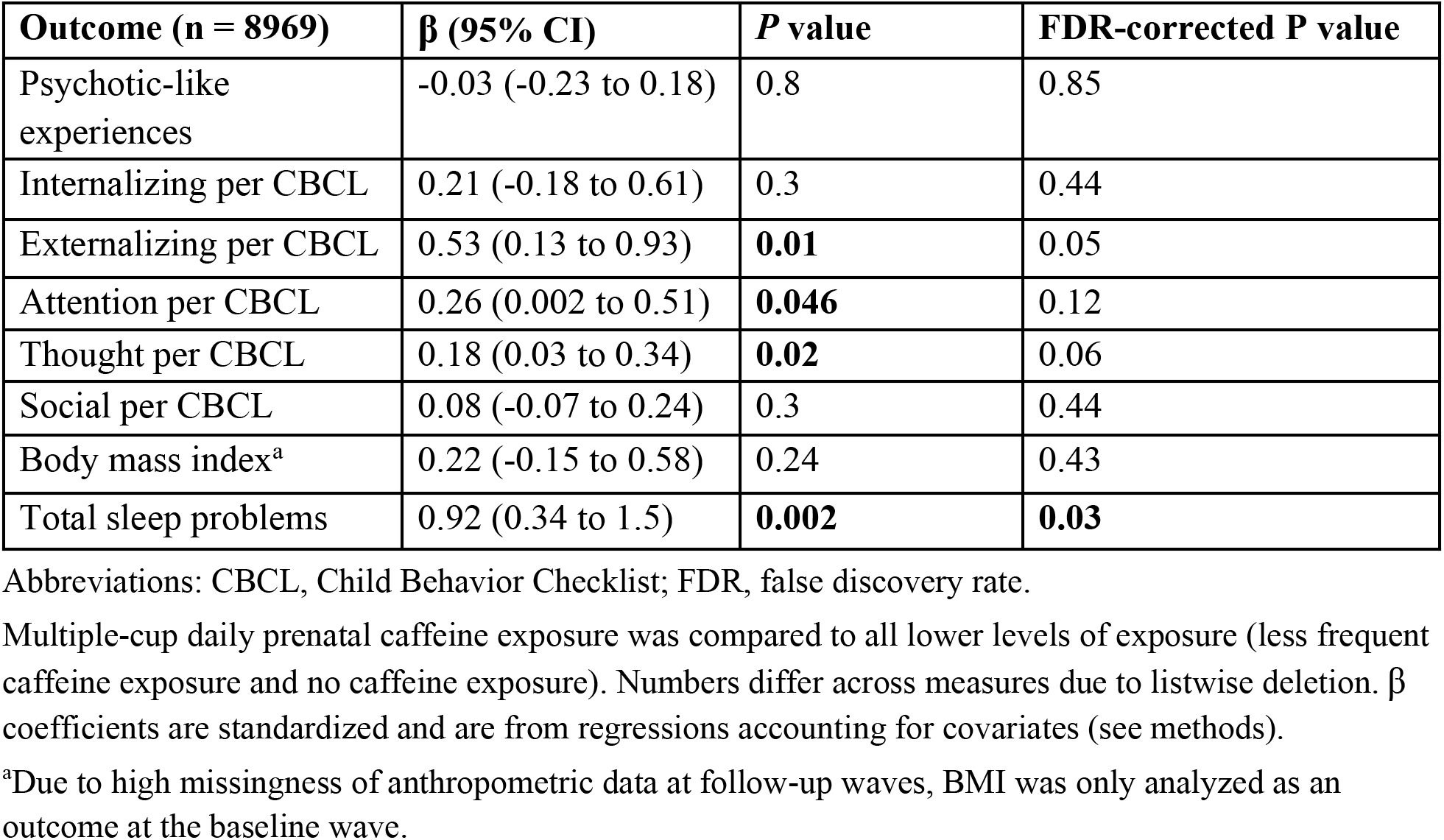
Multiple-Cup Daily Exposure and Child Outcomes.

| **Outcome (n = 8969)** | **$\beta$ (95% CI)** | ***P* value** | **FDR-corrected *P* value** |
| --- | --- | --- | --- |
| Psychotic-like experiences | -0.03 (-0.23 to 0.18) | 0.8 | 0.85 |
| Internalizing per CBCL | 0.21 (-0.18 to 0.61) | 0.3 | 0.44 |
| Externalizing per CBCL | 0.53 (0.13 to 0.93) | **0.01** | 0.05 |
| Attention per CBCL | 0.26 (0.002 to 0.51) | **0.046** | 0.12 |
| Thought per CBCL | 0.18 (0.03 to 0.34) | **0.02** | 0.06 |
| Social per CBCL | 0.08 (-0.07 to 0.24) | 0.3 | 0.44 |
| Body mass index<sup>a</sup> | 0.22 (-0.15 to 0.58) | 0.24 | 0.43 |
| Total sleep problems | 0.92 (0.34 to 1.5) | **0.002** | **0.03** |

Abbreviations: CBCL, Child Behavior Checklist; FDR, false discovery rate.
Multiple-cup daily prenatal caffeine exposure was compared to all lower levels of exposure (less frequent caffeine exposure and no caffeine exposure). Numbers differ across measures due to listwise deletion. $\beta$ coefficients are standardized and are from regressions accounting for covariates (see methods).
<sup>a</sup>Due to high missingness of anthropometric data at follow-up waves, BMI was only analyzed as an outcome at the baseline wave.

**Figure 1.**
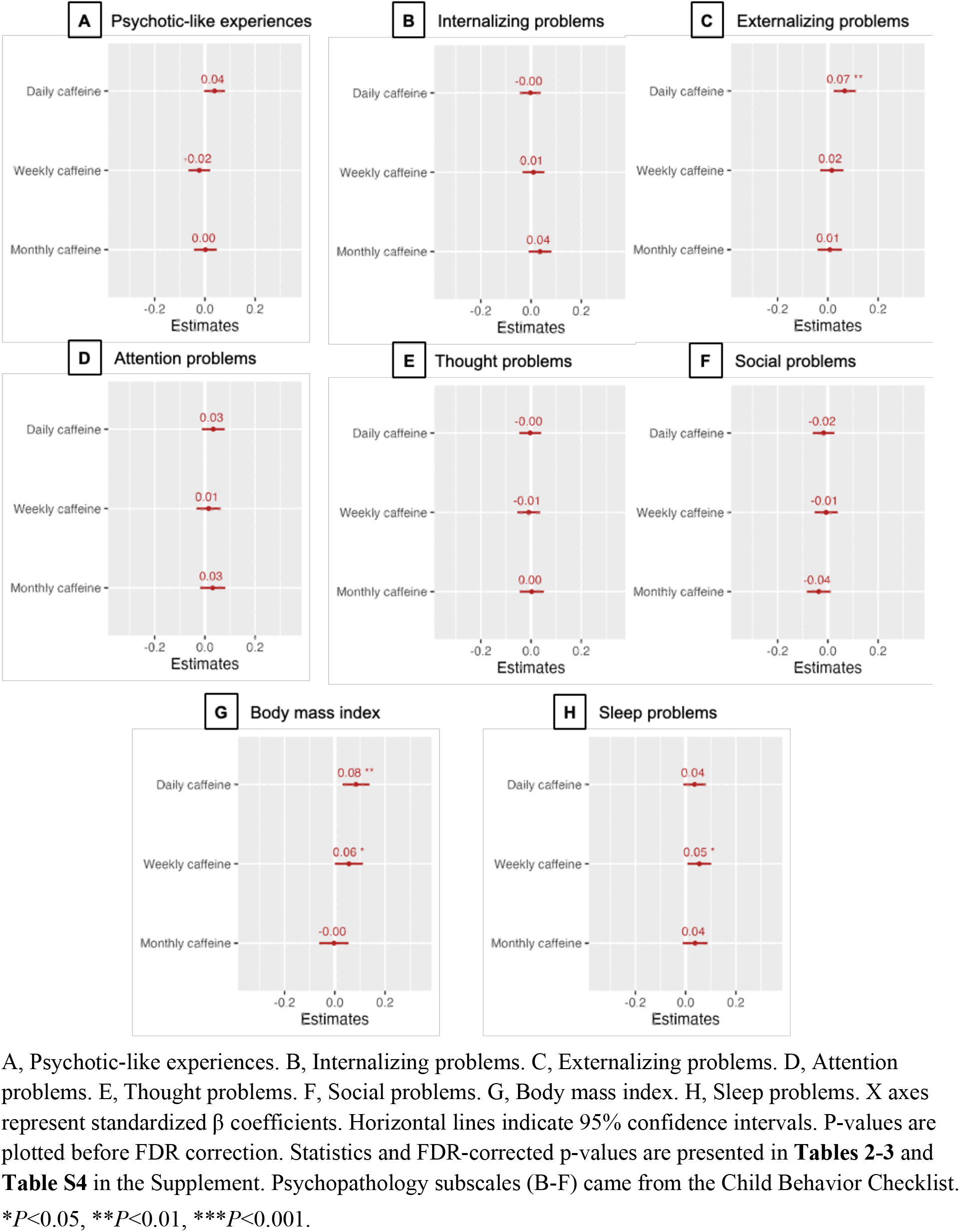
Association of Multiple Levels of Prenatal Caffeine Exposure with Risk of Adverse Childhood Outcomes. A, Psychotic-like experiences. B, Internalizing problems. C, Externalizing problems. D, Attention problems. E, Thought problems. F, Social problems. G, Body mass index. H, Sleep problems. X axes represent standardized β coefficients. Horizontal lines indicate 95% confidence intervals. P-values are plotted before FDR correction. Statistics and FDR-corrected p-values are presented in **Tables 2-3** and **Table S4** in the Supplement. Psychopathology subscales (B-F) came from the Child Behavior Checklist. \**P*<0.05, \*\**P*<0.01, \*\*\**P*<0.001.

#### Child Behavior Problems

Omnibus tests revealed no significant associations between prenatal caffeine exposure and child psychopathology after accounting for multiple testing (all χ^2^<7.19; *ps*>0.07, FDR-corrected *p*>0.15; **Table S1**). There was a nominally significant association between prenatal caffeine exposure and higher externalizing behavior that was not robust to multiple testing correction (χ^2^=10.03; *p*=0.02, FDR-corrected *p*=0.06; **Table S1**). Post hoc group comparisons of this nominally significant omnibus test revealed that this result was driven exclusively by daily caffeine exposure, which was associated with more externalizing problems than less frequent and no exposure (β>0.06; FDR-corrected *p*<0.02; **Table S4**). Any level of caffeine exposure contrasted with no exposure resulted in no significant associations with behavioral problems (all |β|s<0.25; all ps > 0.11; all FDR-corrected *ps*>0.25; **Table S4**). Contrasting those exposed to ≥2 cups/day to those with ≤1 cup/day (i.e., 1 cup/day, weekly, monthly exposure and no exposure) revealed that daily multiple cup caffeine exposure was associated with more sleep problems (β=0.92; FDR-corrected *p*=0.03; **Table 3**). Nominally significant associations that were not robust to FDR correction were also observed for greater externalizing, attention, and thought problems among those with multiple cup exposure (all βs > 0.18; all *ps* < 0.05; all FDR-corrected *ps* < 0.13; **Table 3**).

Age interaction effects were not significantly associated with child behavioral problems after multiple testing correction for all outcomes, with the exception of attention problems (χ^2^=22.82; FDR-corrected *p*=0.01; **Table S2**). Post hoc testing revealed that the age^2^ x prenatal caffeine exposure interaction association with attention problems was attributable to significant differences in age-related quadratic slopes between the monthly and no exposure groups only (β = -0.04; FDR-corrected *p*=0.004; **Table S3**; all other group contrasts: all |β|s < 0.06; all *ps* > 0.01; all FDR-corrected *ps* > 0.05; **Figure S2**; **Table S3**).

### Sensitivity Analyses

Post hoc analyses excluding children who engaged in uncommon substance use, who were exposed to other illicit substances prenatally, who were born at extreme levels of prematurity (**Supp. 1**), or whose biological mothers were not the parent or caregiver respondent revealed associations consistent with those reported above (**Tables S5-S8**).

## Discussion

Our study of prenatal caffeine exposure in the ABCD Study revealed two primary findings. First, daily prenatal caffeine exposure was associated with greater risk for heightened childhood BMI than monthly and no exposure at a small effect size (βs>0.08). Second, we found limited evidence that prenatal caffeine exposure, particularly at low levels, is associated with child behavior problems. Indeed, there was only nominally significant evidence of increased externalizing behavior among those with daily prenatal exposure. However, there were associations of large effects (β=0.92) between multiple cup daily exposure and heightened sleep problems relative to those with less and no exposure. Multiple cup exposure was also associated with higher externalizing, attention, and thought problems at nominal levels of significance that were not robust to multiple testing correction. Collectively, our findings suggest that prenatal caffeine exposure, particularly when it is greater than 1 cup daily, is associated with a small increase in risk for elevated BMI and more sleep problems during childhood/adolescence. Alongside non-human animal experiments showing that prenatal caffeine exposure can influence brain and behavioral development^11,25^, these associations, which were robust to the inclusion of many potentially confounding pregnancy, familial, and child factors, increase the plausibility that prenatal caffeine exposure may have a small impact on childhood BMI and sleep in childhood/adolescence.

### BMI

In contrast to evidence that caffeine use in adulthood is associated with smaller BMI^25^, accumulating evidence supported by meta-analyses and large scale prospective studies have shown that prenatal caffeine exposure is associated with increased weight gain and fat deposition during childhood as well as higher childhood BMI and obesity^26–29^. Indeed, in a prior analysis of ABCD Study data, Zhang and colleagues^14^ found that only high levels of prenatal caffeine exposure (i.e., ≥3 cups) were associated with higher BMI. That we found that any daily exposure was associated with elevated BMI may be attributable to our use of the larger ABCD sample (not contingent upon neuroimaging data) and our categorical comparison of daily exposure with no exposure as opposed to Zhang et al., who evaluated exposure to 3 or more cups and compared this to no exposure. The mechanisms through which prenatal caffeine exposure may contribute to elevated BMI during childhood are unclear. Experimental studies in rodents as well as human association studies have also revealed that prenatal caffeine exposure is associated with lower birth weight and being small-for gestational-age, which have been associated with higher risk of child obesity^29–31^. It is plausible that modifications to the fetal environment (e.g., intrauterine growth restriction)^32^, induced metabolic dysfunction (e.g., hypothalamic-pituitary-adrenal axis function)^33^, and more indirect pathways (e.g., neural reward sensitivity)^34^ may contribute to the development of greater BMI during childhood.

### Child/Adolescent Behavior Problems

Associations between prenatal caffeine exposure and childhood behavioral outcomes have been less studied than BMI and have resulted in an equivocal literature^35,36^. Among child behavior problems examined, we found evidence for elevated sleep problems among children who were prenatally exposed to multiple cups daily. Notably, this association was robust to confounders including child caffeine use. The limited prior studies reporting on sleep in the context of prenatal exposure have predominantly reported null associations. For instance, among 885 infants born, high caffeine consumption was not associated with nighttime infant waking^37^. Further, a prior ABCD Study revealed no association between prenatal caffeine exposure and sleep problems^14^. Similar to the divergence noted above for BMI, this may be attributable to differences in sample size and caffeine groupings (i.e., our study grouping 2 or more cups into multigroup exposure and Zhang et al. using a definition of 3 or more cups). There is a mixed literature surrounding associations between prenatal caffeine exposure and childhood externalizing with the majority of studies finding no evidence of association^35,38^. Notably, like a prior report arising from the ABCD Study finding^14^, our study revealed that daily and daily multiple cup prenatal caffeine exposure were nominally significantly associated with higher externalizing problems, but were not robust to multiple testing correction.

The observed association with sleep problems may have multiple mechanisms. Caffeine is an adenosine receptor antagonist involved in numerous intracellular signaling pathways^39^. At high doses, it may interfere with ɣ-amino butyric acid (GABA) receptors^40^. While these concentrations are rarely seen in mild to moderate caffeine consumers, the decreased ability to clear caffeine during pregnancy as well as its extended half-life within the fetus may cause the substance to accumulate to levels that could impact GABA function^3^. Indeed, altered GABA neuron migration during development has been observed among mouse pups with caffeine exposure during gestation^11^. These alterations may affect the excitatory-inhibitory balance of the brain, which must be tightly regulated to maintain the sleep-wake cycle^41^. However, it is also plausible that our observed association between daily multiple cup caffeine exposure during prenatal development and sleep problems reflects epiphenomena. For instance, caffeine consumption is associated with reductions in total sleep and less sleep during pregnancy is associated with greater sleep problems and others (e.g., externalizing) among offspring^42,43^. Thus it is possible that these associations arise not as a direct consequence of caffeine but on its impact on maternal sleep during pregnancy or a multitude of other factors.

Finally, our longitudinal analyses revealed that attention problems were associated with a significant interaction between prenatal caffeine exposure and age. Post-hoc analyses found that this effect was driven by the interaction between monthly prenatal caffeine exposure (as compared to no exposure) and age^2^, where-in the expected age-associated decrease in attention problems^44^ was delayed in the monthly-exposure group (**Figure S2**). While further details on reasons for caffeine use or on use patterns were not available (e.g., the total number of drinks consumed in one drinking period), the specificity of the effect suggests that the monthly use group may be heterogeneous. For example, differences in caffeine sensitivity, use patterns, and motivations for use could be driving results (e.g., infrequent binges during periods of high stress). It may be fruitful for future work to explore motivations for and patterns of use with greater detail, when testing associations with prenatal caffeine exposure.

### Limitations

It is important to interpret results in the context of limitations. First, caffeine consumption during pregnancy was assessed using retrospective caregiver reports which may introduce misclassification resulting from memory errors and/or social desirability. For example, one study comparing mothers’ retrospective reporting of prenatal experiences at 6-months postpartum and at 8-years postpartum found less agreement in evaluation of prescription usage (“doctor recommended pills”) than in other antenatal events^45^. With that said, the caffeine use observed in our study (i.e., 60% of mothers consuming caffeine overall and 25% consuming caffeine daily) aligns with other estimates from national datasets^1,46^. Second, caffeine dosage was measured using unspecified cups per day, which does not take into account the variable amounts of caffeine in different caffeinated drinks or other sources. Third, rodent studies have shown that caffeine consumption specifically during mid-late pregnancy may result in adverse effects on the fetal neuroendocrine system^5^, but the ABCD dataset does not contain information on when during the gestational period the participants were exposed. As further data from this cohort are released, it would be important to retest associations with prenatal caffeine exposure as children age into middle and late adolescence. Finally, it remains possible that factors we were not able to account for that may be correlated with caffeine consumption during pregnancy including other lifestyle factors (e.g., other nutrition) and experiences (e.g., stress, anxiety), and correlates of caffeine being in food (e.g., sugar, sweeteners) may have contributed to observed associations. However, experimental caffeine administration in non-human animal models have found data consistent with these associations^11,26^. Nonetheless, despite these limitations, our findings align with evidence from large prospective cohorts linking prenatal caffeine exposure to heightened BMI and are consistent with non-human studies showing that prenatal caffeine exposure contributes to factors associated with later childhood obesity^28^.

### Conclusion

Limitations notwithstanding, we find that daily caffeine consumption during pregnancy, particularly when occurring more than once, is associated with increased risk for heightened childhood BMI and childhood/adolescent sleep problems. Our observed associations do not indicate whether prenatal caffeine exposure causally contributes to these adverse physical and behavioral outcomes. However, as these associations were robust to many potential confounds and are consistent with prospective studies in humans and experimental models in non-human animals^34,43^, it remains plausible that prenatal caffeine exposure may contribute to these adverse outcomes. Alongside the need for more studies, and in particular studies of plausible mechanisms, these data suggest that caffeine use during pregnancy should be approached with caution.

## Supporting information

Supplemental Results

## Data Availability

All data produced in the present work are contained in the manuscript.

## Acknowledgements

Data used in the preparation of this article were obtained from the Adolescent Brain Cognitive Development (ABCD) Study (https://abcdstudy.org), held in the NIMH Data Archive (NDA). This is a multisite, longitudinal study designed to recruit more than 10,000 children aged 9 to 10 years and follow them over 10 years into early adulthood. The ABCD Study is supported by the National Institutes of Health. A full list of supporters is available at https://abcdstudy.org/federal-partners.html. A listing of participating sites and a complete listing of the study investigators can be found at https://abcdstudy.org/consortium_members/. ABCD consortium investigators designed and implemented the study and/or provided data but did not necessarily participate in analysis or writing of this report. The ABCD data repository grows and changes over time. The ABCD data used in this report came from 10.15154/8873-zj65. The DOIs can be found at http://dx.doi.org/10.15154/8873-zj65. This study was funded by R01DA054750 (RB, AA). Authors received additional funding support from the National Institutes of Health: SEP (F31AA029934), AJG (NSF DGE-213989), DAAB (K99AA030808), JB (R01MH128286, R01MH132962), and RB (R21AA027827, R01HD113188, R01AG061162, U01DA055367, U01DA055367S)

