## Supplemental Methods for "Associations between prenatal caffeine exposure and child development: Longitudinal results from the Adolescent Brain Cognitive Development (ABCD) Study"

**Supplement 1: Methods.**

**MEASURES**

**Psychotic-Like Experiences.** On the 21-item Prodromal Questionnaire-Brief Child Version (PQ-BC)[1], children are asked to respond (yes/no) to whether they experienced a thought/feeling/experience (e.g., do familiar surroundings sometimes seem strange, confusing, threatening, or unreal to you?). They then report on whether the event was distressing and if so, the extent that it bothered them. Total scores were derived from the sum of the endorsed items. In this sample, the winsorized total scores (±3 SDs) ranged from 0-9 (non-winsorized, 0-21).

**CBCL Internalizing and Externalizing Subfacets.** The 113-item Child Behavior Checklist (CBCL)[2] was completed by parents to evaluate specific problems that their children encountered within the last six months. They rated these items on a scale from 0 (*not true [as far as you know]*) to 2 (*very true or often true*). The Internalizing problems scale consists of three subscales, which examine *anxious/depressed*, *withdrawn/depressed*, and *somatic* problems. Likewise, the CBCL Externalizing problems scale comprises subscales for *rule-breaking* and *aggressive behavior*. In this sample, winsorized internalizing scores ranged from 0-16 (non-winsorized, 0-51) and externalizing from 0-15 (non-winsorized, 0-50).

**Sleep Problems.** The Sleep Disturbance Scale for Children (SDSC)[10] is a questionnaire consisting of 27 items, which parents complete to evaluate sleep problems in their children. The total score is calculated by adding up the scores from six subscales that assess specific symptoms: 1) disorders of initiating and maintaining sleep, 2) sleep breathing disorders, 3) disorders of arousal, 4) sleep-wake transition disorders, 5) disorders of excessive somnolence, and 6) sleep hyperhidrosis. The total score is obtained by summing the scores from these six subscales. In this sample, the winsorized total sleep score ranged from 28 to 52 (non-winsorized, 26-126).

**Body Mass Index (BMI).** BMI was measured in children at baseline using the following formula:

$$703 * \frac{weight\ (lbs)}{height\ (in)^2}$$

Data with implausible values were set to missing in our analyses according to the following criteria. First, BMI measures based on impossible values (e.g., height of 4 inches) were set to missing (n=8). Second, BMIs outside of the range of CDC charts[11] (i.e. 10) were set to missing (n=101). Overall, follow-up BMI data points were removed due to missing height or weight requirements (~25%), with 109 BMI values set to missing based on the aforementioned data cleaning. In this sample, winsorized values ranged from 14.3-27.1 (non-winsorized, 12.0-55.0).

**Covariate Descriptions**

All variables were assessed at the ABCD baseline assessment and 1/2/3 year follow-ups with the exception of prenatal exposure, birth weight, and gestational age at birth reflect measures taken while children were ages 9.9±0.6 years.

**Child age.** Age was self-reported in months and converted to years.

**Child sex.** Child sex is coded as a dichotomous variable indicating whether the child is male (1) or female (2).

**Birth weight.** Parents/caregivers retrospectively reported on child birth weight and pounds and ounces, which we converted to total ounces.

**Child Race/Ethnicity.** Parents/caregivers selected from 26 different categories. We formed dichotomous groups for the most prevalent categories of race (i.e., White, Black, Asian, Pacific Islander, Native American), with remaining participants being assigned to Other. Hispanic ethnicity was also included. All variables were dummy coded as non-mutually exclusive dichotomous variables; as such, participants could be coded within more than one category.

**Child Lifetime Substance Exposure.** Children self-reported if they tried alcohol (i.e., ever had a sip), marijuana (e.g. ever had a puff of a cigarette or blunt, etc.), and tobacco (e.g., ever had a puff of a cigarette or hookah, etc.). More substantive self-reported use or positive hair toxicology screens were rare and as such participants were excluded on these metrics in follow-up analyses (for additional information on these variables, please refer to the section entitled Post-hoc Analyses with Exclusions: Excluding Uncommon Substances Used by Children and Mothers During Pregnancy as well as Children Born Prematurely and non-Biological Mother Report below).

**Child Caffeine Summary Score.** Children self-reported the number of drinks of coffee, espresso, tea, soda, and energy drinks they consumed in the past six months. These numbers were added to form a caffeine summary score. In this sample, winsorized values ranged from 0-8 (non-winsorized, 0-336).

**Unplanned Pregnancy.** Parents/caregivers retrospectively reported whether the pregnancy was planned.

**Maternal Age at Birth.** Parents/caregivers retrospectively reported the child's biological mother's age at the time of the child's birth.

**Prenatal Vitamin Usage.** Parents/caregivers noted whether the mother used prenatal vitamins during the pregnancy.

**Gestational Age at Birth.** Parents/caregivers retrospectively reported on child gestational age at birth.

**Gestational Age when Mother Learned of Pregnancy.** Parents/caregivers retrospectively reported the number of weeks the mother was pregnant before she learned of her pregnancy.

**Maternal Education.** Maternal education was recoded such that 12th grade, HS grad, and GED=12 years; some college and associate's degree = 14 years; Bachelor's degree = 16 years; Master's degree = 18 years; Professional and Doctoral degrees = 20 years.

**Household Income.** Due to low endorsement of the first five of 10 household income levels, this variable was recoded such that the first five categories were assigned a value of one (i.e., <$50,000; n=2,914). The subsequent categories used were coded as two ($50,000-$74,999, n=1,385), three ($75,000-$99,999, n=1,439), four ($100,000-$199,999, n=3,069), and five ($200,000 or more, n=1,173), respectively (**Table 1**).

**Family History of Psychopathology.** Parents/caregivers reported on whether any 1st degree relatives had a history of problems with psychosis, depression, anxiety, antisocial behavior, or mania.

**Adult Self Report.** Primary parents reported on their own adaptive functioning, problems, and substance use using the 126-item Adult Self Report checklist[12]. Baseline values for behavioral subscales corresponding to CBCL subscales of interest (internalizing, externalizing, attention problems, thought problems) were included as covariates in the model.

**Prenatal Exposure to alcohol, tobacco, and cannabis.** Child prenatal exposure to alcohol, tobacco, and/or cannabis both before (alcohol: n = 2,698; tobacco: n = 1,465; cannabis: n = 606) and after (alcohol: n = 274; tobacco: n = 554; cannabis: n = 209) maternal knowledge of pregnancy were coded as separate dichotomous variables based upon parent/caregiver retrospective report.

**Post-hoc Analyses with Exclusions: Uncommon Substances Used by Children and Mothers During Pregnancy as well as Children Born Prematurely and non-Biological Mother Report.** A series of post hoc analyses were conducted to determine whether removing the following individuals from analyses significantly altered results:

a) children who reported using marijuana (n=72), using substances other than alcohol and tobacco (i.e., bath salts, n=2; "sniff" to "get high," n=9; inhalants, n=14; amphetamines not as prescribed, n=9; tranquilizers/anxiolytics/sedatives not as prescribed, n=11; pain medication not as prescribed, n=4; cough medication to "get high," n=8), or having a full drink of alcohol (n=123) or more than a puff of tobacco (n=36), or screened positive for substances based on hair toxicology (methamphetamines, n=11; THC, n=95; EtG, n=37; cotinine, n=9; Adderall, n=82). Individuals were removed if they reported using the above substances at any of the timepoints used in the analysis (baseline to 3YFU). A total of 409 individuals were excluded for these analyses (Numbers do not sum as some children were positive across multiple indices).

b) individuals whose biological mothers reported using other illicit substances while pregnant (cocaine or crack, n=51; heroin or morphine, n=19; oxycontin, n=25; other, n=33). A total of 102 individuals were excluded (numbers do not sum as some mothers were positive across multiple indices).

c) individuals born more than 8 weeks premature (n=149).

d) individuals whose parent/caregiver in ABCD is not the biological mother (n=1617).

**Supplemental References.**


1. Loewy RL, Pearson R, Vinogradov S, Bearden CE, Cannon TD. Psychosis risk screening with the Prodromal Questionnaire — Brief Version (PQ-B). *Schizophr Res*. 2011;129(1):42-46. doi:10.1016/j.schres.2011.03.029
2. Achenbach TM, Rescorla L. *Manual for the ASEBA School-Age Forms & Profiles: An Integrated System of Multi-Informant Assessment*. ASEBA; 2001.
3. Akshoomoff N, Beaumont JL, Bauer PJ, et al. VIII. NIH TOOLBOX COGNITION BATTERY (CB): COMPOSITE SCORES OF CRYSTALLIZED, FLUID, AND OVERALL COGNITION: NIH TOOLBOX COGNITION BATTERY (CB). *Monogr Soc Res Child Dev*. 2013;78(4):119-132. doi:10.1111/mono.12038
4. Eriksen BA, Eriksen CW. Effects of noise letters upon the identification of a target letter in a nonsearch task. *Percept Psychophys*. 1974;16(1):143-149. doi:10.3758/BF03203267
5. Tulsky DS, Carlozzi NE, Chevalier N, Espy KA, Beaumont JL, Mungas D. V. NIH TOOLBOX COGNITION BATTERY (CB): MEASURING WORKING MEMORY: NIH TOOLBOX COGNITION BATTERY (CB). *Monogr Soc Res Child Dev*. 2013;78(4):70-87. doi:10.1111/mono.12035
6. Zelazo PD. The Dimensional Change Card Sort (DCCS): a method of assessing executive function in children. *Nat Protoc*. 2006;1(1):297-301. doi:10.1038/nprot.2006.46
7. Dikmen SS, Bauer PJ, Weintraub S, et al. Measuring Episodic Memory Across the Lifespan: NIH Toolbox Picture Sequence Memory Test. *J Int Neuropsychol Soc*. 2014;20(6):611-619. doi:10.1017/S1355617714000460
8. Carlozzi NE, Tulsky DS, Kail RV, Beaumont JL. VI. NIH TOOLBOX COGNITION BATTERY (CB): MEASURING PROCESSING SPEED: NIH TOOLBOX COGNITION BATTERY (CB). *Monogr Soc Res Child Dev*. 2013;78(4):88-102. doi:10.1111/mono.12036
9. Gershon RC, Slotkin J, Manly JJ, et al. IV. NIH TOOLBOX COGNITION BATTERY (CB): MEASURING LANGUAGE (VOCABULARY COMPREHENSION AND READING DECODING): NIH TOOLBOX COGNITION BATTERY (CB). *Monogr Soc Res Child Dev*. 2013;78(4):49-69. doi:10.1111/mono.12034
10. Bruni O, Ottaviano S, Guidetti V, et al. The Sleep Disturbance Scale for Children (SDSC) Construct ion and validation of an instrument to evaluate sleep disturbances in childhood and adolescence. *J Sleep Res*. 1996;5(4):251-261. doi:10.1111/j.1365-2869.1996.00251.x
11. Kuczmarski RJ, Ogden CL, Guo SS, et al. 2000 CDC Growth Charts for the United States: methods and development. *Vital Health Stat 11*. 2002;(246):1-190.
12. Achenbach TM, Rescorla L. *Manual for the ASEBA Adult Forms & Profiles: For Ages 18-59: Adult Self-Report, Adult Behavior Checklist*. ASEBA; 2003.
