## Supplemental Results for "Associations between prenatal caffeine exposure and child development: Longitudinal results from the Adolescent Brain Cognitive Development (ABCD) Study"

**Supplement 2: Results**

**Longitudinal Outcomes of Exposure Without Covariates**

Before covariate adjustment, any level of prenatal caffeine exposure was associated with social, attention, thought, internalizing, externalizing, and sleep problems (all $\beta > 0.13$; all FDR-corrected $P < .008$; **eTable 1**). Daily exposure was associated with higher scores on these scales as well, in addition to increased PLEs (all $\beta > 0.17$; all FDR-corrected $P < .04$; **eTable 1**).

**Outcomes of Exposure Without Covariates in the Daily Caffeine Subset**

Longitudinal analysis without covariate adjustment in all daily users showed that multiple-cup daily caffeine exposure was associated with increased scores on all five CBCL subscales, as well as increased PLEs and sleep problems ($\beta > 0.11$, FDR-corrected $P < .02$; **eTable 2**). Any daily exposure as compared to controls was also marginally/nominally significant for an increase in attention problems (($\beta = 0.1$; $P = .05$; FDR-corrected $P = 0.2$; **eTable 2**).

**Analyses with Exclusions.**

**Excluding children for uncommon substance use.** Results were largely consistent with those reported using the full sample before FDR correction (**eTables 3 and 7**), the only exceptions being that children exposed to multiple cups of caffeine daily had significantly higher social and thought problems than those exposed to one cup or less daily (all $\beta > 0.154$, all $p < 0.05$), and the significant difference in attention problems between those exposed daily and those exposed less frequently was reduced to a trend ($\beta = 0.142$, $p = 0.071$).

**Excluding children exposed to uncommon substances during pregnancy.** Results were consistent with those reported using the full sample (**eTables 4 and 8**).

**Excluding children born prematurely.** Results were consistent with those reported using the full sample (**eTables 5 and 9**).

**Excluding children whose parent/caregiver report was not their biological mother.** As described in **eTables 6 and 10**, results were consistent with those reported using the full sample.
