## Supplemental Figure 1 for "Associations between prenatal caffeine exposure and child development: Longitudinal results from the Adolescent Brain Cognitive Development (ABCD) Study"

**Figure S1.** Distribution of Prenatal Caffeine Exposure Across Cohort.

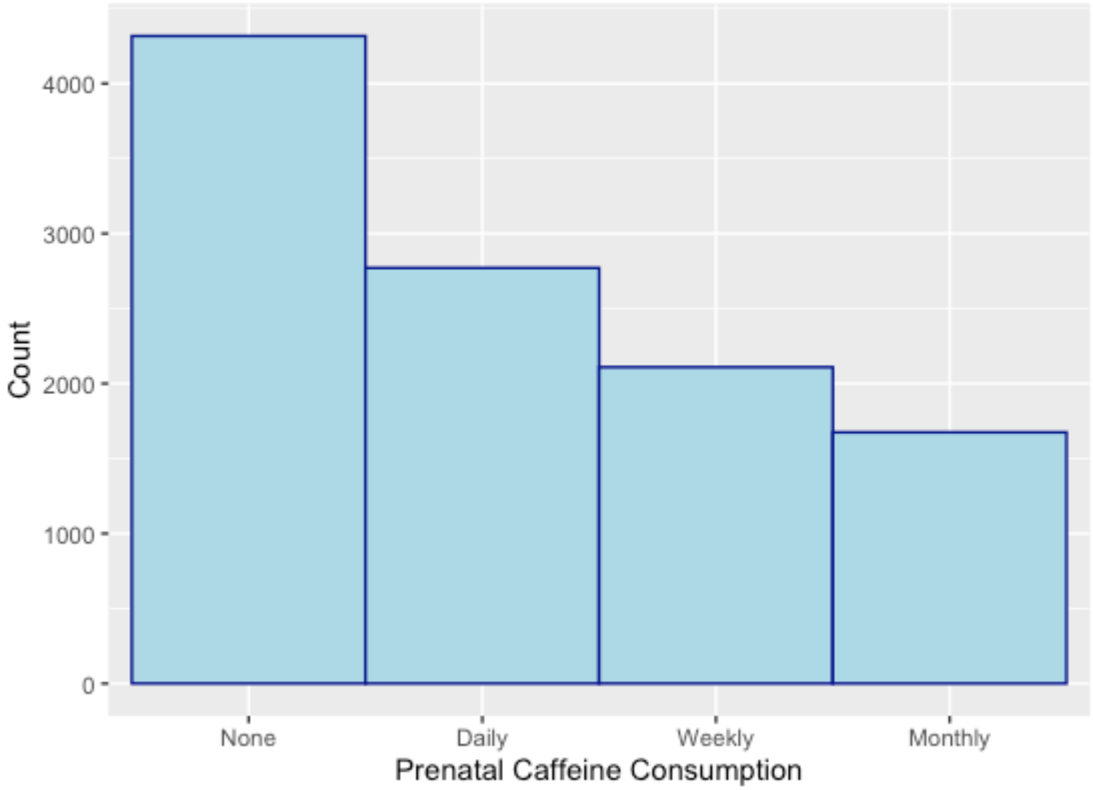


***Figure S1 Note.*** Prenatal caffeine consumption was coded within the ABCD dataset as a categorical variable with four levels: none, daily, weekly, and monthly exposure. Maternal caffeine consumption during pregnancy was a retrospective self-report taken at the baseline timepoint. Exact count in each group is presented in **Table 1**.
