## Supplemental Figure 2 for "Associations between prenatal caffeine exposure and child development: Longitudinal results from the Adolescent Brain Cognitive Development (ABCD) Study"

**Figure S2.** Age Interaction Plot for Prenatal Caffeine Exposure and Attention Problems.

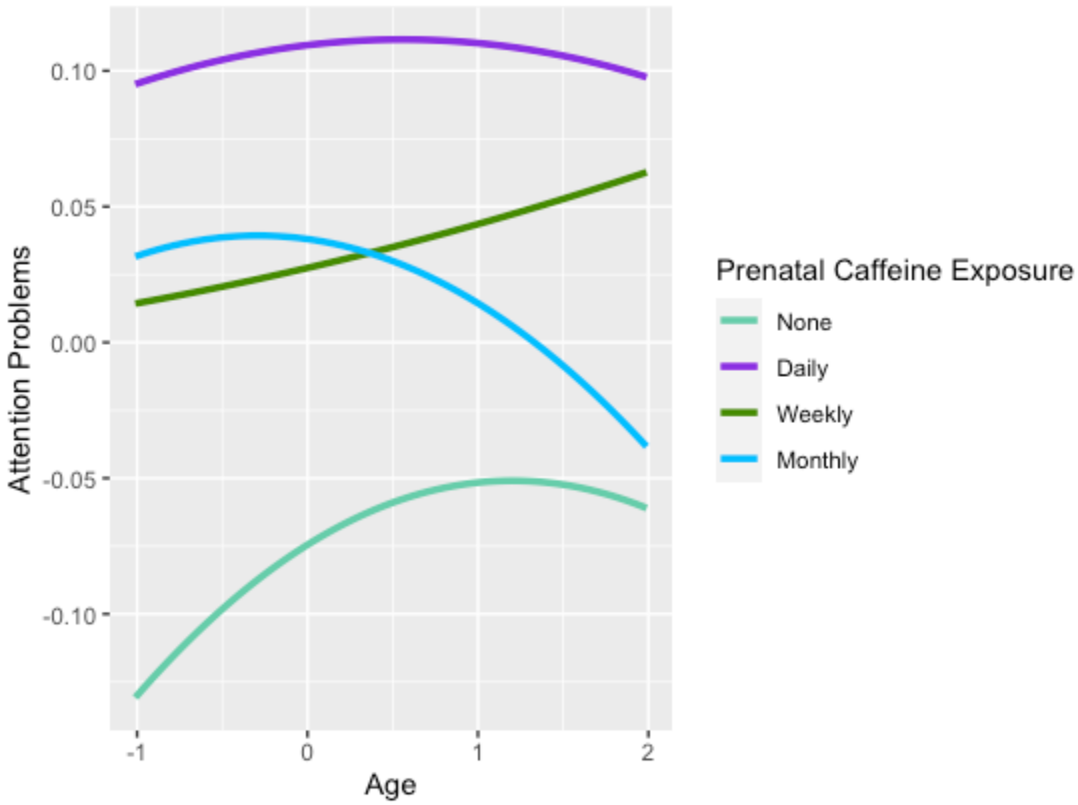


***Figure S2 Note.*** Age effects were determined using a linear mixed-effects model. Age and age squared values were derived from the ABCD interview age variable. Attention problems were derived from the Child Behavior Checklist. Both axes were log-transformed and normalized. Prenatal caffeine consumption was coded within the ABCD dataset as a four-level categorical variable.
