## Supplemental Table 1 for "Associations between prenatal caffeine exposure and child development: Longitudinal results from the Adolescent Brain Cognitive Development (ABCD) Study"

**Table S1.** Prenatal Caffeine Exposure and Child Outcomes Omnibus Testing

| **Outcome (n = 8969)** | $\chi^2$ | ***P* value** | **FDR-corrected P value** |
|---|---|---|---|
| Psychotic-like experiences | 7.19 | 0.07 | 0.19 |
| Internalizing per CBCL | 2.99 | 0.39 | 0.53 |
| Externalizing per CBCL | 10.03 | **0.02** | 0.08 |
| Attention per CBCL | 2.81 | 0.42 | 0.53 |
| Thought per CBCL | 0.22 | 0.98 | 0.98 |
| Social per CBCL | 2.57 | 0.46 | 0.53 |
| Body mass index[b] | 12.57 | **5.67E-03** | **0.045** |
| Total sleep problems | 5.98 | 0.11 | 0.22 |

***Table S1 Note.*** Log-likelihood ratio tests were used to analyze the associations between prenatal caffeine exposure (a factor variable with 4 mutually exclusive groups: no, daily, weekly, and monthly exposure) and each outcome, nesting data by research site and family identification. All covariates listed in the methods were included in the analysis. Psychotic-like experiences were assessed with the Prodromal Questionnaire Brief-Report Child Version and sleep problems from the Parent Sleep Disturbance Scale for Children.

[a]Due to high missingness of anthropometric data at follow-up waves, BMI was only analyzed as an outcome at the baseline wave.
