## Supplemental Table 2 for "Associations between prenatal caffeine exposure and child development: Longitudinal results from the Adolescent Brain Cognitive Development (ABCD) Study"

**Table S2.** Age Interaction Effects on Associations Between Prenatal Caffeine Exposure and Outcomes of Interest

| **Outcome** | $\chi^2$ | ***P* value** | **FDR-corrected P value** |
|---|---|---|---|
| Psychotic-like experiences | 2.47 | 0.87 | 0.87 |
| Internalizing per CBCL | 9.64 | 0.14 | 0.25 |
| Externalizing per CBCL | 12.32 | 0.06 | 0.19 |
| Attention per CBCL | 22.82 | **8.58E-04** | **0.01** |
| Thought per CBCL | 4.14 | 0.66 | 0.77 |
| Social per CBCL | 11.02 | 0.09 | 0.21 |
| Total sleep problems | 6.82 | 0.34 | 0.47 |

***Table S2 Note.*** Prenatal caffeine exposure was coded as a 4-level categorical variable in the ABCD dataset (daily, weekly, monthly, and no exposure). Two separate linear mixed-effects models were used to analyze age effects: 1) interview age included as a covariate; and 2) age x caffeine and $age^2$ x caffeine included as interaction terms. These models were compared using log-likelihood ratio tests to determine the outcomes with significant age interactions.
