## Supplemental Table 3 for "Associations between prenatal caffeine exposure and child development: Longitudinal results from the Adolescent Brain Cognitive Development (ABCD) Study"

**Table S3.** Post Hoc Analysis of Age Effects on Attention Problems and Prenatal Caffeine Exposure Associations

| | Age | | | $Age^2$ | | |
|---|---|---|---|---|---|---|
| **Contrast** | **β** | ***P* value** | **FDR-corrected P value** | **β** | ***P* value** | **FDR-corrected P value** |
| Daily vs. no exposure | -0.01 | 0.41 | 0.55 | -0.02 | **0.02** | 0.06 |
| Weekly vs. no exposure | 0.02 | **0.04** | 0.08 | -0.01 | 0.45 | 0.52 |
| Monthly vs. no exposure | -0.01 | 0.54 | 0.61 | -0.04 | **4.38E-04** | **0.004** |
| Daily vs. weekly | -0.03 | **0.01** | 0.07 | -0.01 | 0.2 | 0.27 |
| Daily vs. monthly | -0.001 | 0.93 | 0.93 | 0.02 | 0.16 | 0.25 |
| Weekly vs. monthly | 0.03 | **0.02** | 0.07 | 0.03 | **0.01** | 0.05 |
| Any caffeine exposure vs. no exposure | 0.06 | **0.02** | 0.07 | -0.04 | 0.07 | 0.15 |
| Daily caffeine exposure vs. all lower exposure | -0.05 | 0.14 | 0.23 | 0.01 | 0.74 | 0.74 |

***Table S3 Note.*** Prenatal caffeine exposure was coded as a 4-level categorical variable in the ABCD dataset (daily, weekly, monthly, and no exposure). Two separate linear mixed-effects models were used to analyze age effects: 1) interview age included as a covariate; and 2) age x caffeine and $age^2$ x caffeine included as interaction terms. β coefficients are standardized and are from regressions accounting for covariates (see methods).
