## Supplemental Table 4 for "Associations between prenatal caffeine exposure and child development: Longitudinal results from the Adolescent Brain Cognitive Development (ABCD) Study"

**Table S4.** Associations of All Outcomes with All Prenatal Caffeine Exposure Levels

| Outcome (n = 8991) | Contrast | β | *P* value | FDR-corrected *p* value |
|---|---|---|---|---|
| **Psychotic-like experiences** | Daily caffeine exposure vs no exposure | 0.04 | 0.07 | 0.21 |
| | Weekly caffeine exposure vs no exposure | -0.02 | 0.31 | 0.49 |
| | Monthly caffeine exposure vs no exposure | 0.002 | 0.93 | 0.93 |
| | Daily caffeine exposure vs weekly exposure | 0.06 | **0.01** | 0.09 |
| | Daily caffeine exposure vs monthly exposure | 0.04 | 0.15 | 0.34 |
| | Weekly caffeine exposure vs monthly exposure | -0.03 | 0.75 | 0.49 |
| | Any caffeine exposure vs no exposure | -0.05 | 0.38 | 0.49 |
| | Daily caffeine exposure vs all lower exposure | 0.14 | **0.02** | 0.13 |
| **Internalizing per CBCL** | Daily caffeine exposure vs no exposure | -0.003 | 0.89 | 0.89 |
| | Weekly caffeine exposure vs no exposure | 0.01 | 0.67 | 0.89 |
| | Monthly caffeine exposure vs no exposure | 0.04 | 0.12 | 0.54 |

| | | | | |
|---|---|---|---|---|
| | Daily caffeine exposure vs weekly exposure | -0.01 | 0.6 | 0.89 |
| | Daily caffeine exposure vs monthly exposure | -0.04 | 0.12 | 0.54 |
| | Weekly caffeine exposure vs monthly exposure | -0.03 | 0.31 | 0.7 |
| | Any caffeine exposure vs no exposure | -0.02 | 0.86 | 0.89 |
| | Daily caffeine exposure vs all lower exposure | -0.05 | 0.71 | 0.89 |
| **Externalizing per CBCL** | Daily caffeine exposure vs no exposure | 0.07 | **0.003** | **0.01** |
| | Weekly caffeine exposure vs no exposure | 0.02 | 0.5 | 0.72 |
| | Monthly caffeine exposure vs no exposure | 0.01 | 0.75 | 0.77 |
| | Daily caffeine exposure vs weekly exposure | 0.05 | **0.04** | 0.08 |
| | Daily caffeine exposure vs monthly exposure | 0.06 | **0.03** | 0.06 |
| | Weekly caffeine exposure vs monthly exposure | 0.01 | 0.77 | 0.77 |

| | | | | |
|---|---|---|---|---|
| | Any caffeine exposure vs no exposure | -0.07 | 0.56 | 0.72 |
| | Daily caffeine exposure vs all lower exposure | 0.39 | **0.002** | **0.01** |
| **Attention per CBCL** | Daily caffeine exposure vs no exposure | 0.03 | 0.15 | 0.51 |
| | Weekly caffeine exposure vs no exposure | 0.02 | 0.54 | 0.74 |
| | Monthly caffeine exposure vs no exposure | 0.03 | 0.23 | 0.51 |
| | Daily caffeine exposure vs weekly exposure | 0.02 | 0.48 | 0.74 |
| | Daily caffeine exposure vs monthly exposure | 0.003 | 0.93 | 0.93 |
| | Weekly caffeine exposure vs monthly exposure | -0.02 | 0.58 | 0.74 |
| | Any caffeine exposure vs no exposure | 0.03 | 0.67 | 0.75 |
| | Daily caffeine exposure vs all lower exposure | 0.1 | 0.21 | 0.51 |
| **Thought per CBCL** | Daily caffeine exposure vs no exposure | -0.004 | 0.88 | 0.92 |

| | | | | |
|---|---|---|---|---|
| | Weekly caffeine exposure vs no exposure | -0.01 | 0.7 | 0.92 |
| | Monthly caffeine exposure vs no exposure | 0.003 | 0.92 | 0.92 |
| | Daily caffeine exposure vs weekly exposure | 0.01 | 0.82 | 0.92 |
| | Daily caffeine exposure vs monthly exposure | -0.01 | 0.82 | 0.92 |
| | Weekly caffeine exposure vs monthly exposure | -0.01 | 0.67 | 0.92 |
| | Any caffeine exposure vs no exposure | -0.03 | 0.46 | 0.92 |
| | Daily caffeine exposure vs all lower exposure | 0.04 | 0.38 | 0.92 |
| **Social per CBCL** | Daily caffeine exposure vs no exposure | -0.02 | 0.45 | 0.68 |
| | Weekly caffeine exposure vs no exposure | -0.01 | 0.77 | 0.77 |
| | Monthly caffeine exposure vs no exposure | -0.04 | 0.13 | 0.6 |
| | Daily caffeine exposure vs weekly exposure | -0.01 | 0.69 | 0.77 |

| | | | | |
|---|---|---|---|---|
| | Daily caffeine exposure vs monthly exposure | 0.02 | 0.45 | 0.68 |
| | Weekly caffeine exposure vs monthly exposure | 0.03 | 0.28 | 0.63 |
| | Any caffeine exposure vs no exposure | -0.07 | 0.11 | 0.6 |
| | Daily caffeine exposure vs all lower exposure | 0.02 | 0.63 | 0.77 |
| **Body mass index** | Daily caffeine exposure vs no exposure | 0.08 | **2.02E-03** | **0.02** |
| | Weekly caffeine exposure vs no exposure | 0.06 | **0.049** | 0.13 |
| | Monthly caffeine exposure vs no exposure | -0.004 | 0.91 | 0.91 |
| | Daily caffeine exposure vs weekly exposure | 0.03 | 0.36 | 0.48 |
| | Daily caffeine exposure vs monthly exposure | 0.09 | **7.99E-03** | **0.03** |
| | Weekly caffeine exposure vs monthly exposure | 0.06 | 0.07 | 0.14 |
| | Any caffeine exposure vs no exposure | 0.16 | 0.11 | 0.18 |

| | | | | |
|---|---|---|---|---|
| | Daily caffeine exposure vs all lower exposure | 0.07 | 0.63 | 0.72 |
| **Total sleep problems** | Daily caffeine exposure vs no exposure | 0.04 | 0.12 | 0.25 |
| | Weekly caffeine exposure vs no exposure | 0.06 | **0.02** | 0.1 |
| | Monthly caffeine exposure vs no exposure | 0.04 | 0.13 | 0.25 |
| | Daily caffeine exposure vs weekly exposure | -0.02 | 0.47 | 0.7 |
| | Daily caffeine exposure vs monthly exposure | -0.002 | 0.95 | 0.95 |
| | Weekly caffeine exposure vs monthly exposure | 0.02 | 0.54 | 0.7 |
| | Any caffeine exposure vs no exposure | 0.24 | 0.14 | 0.25 |
| | Daily caffeine exposure vs all lower exposure | -0.02 | 0.9 | 0.95 |

***Table S4 Note.*** Linear mixed-effects models were used to analyze the associations between prenatal caffeine exposure and each outcome, nesting data by research site and family identification. Psychotic-like experiences were assessed with the Prodromal Questionnaire Brief-Report Child Version and sleep problems from the Parent Sleep Disturbance Scale for Children. The β coefficients are standardized.

FDR multiple testing correction was conducted across contrasts for each outcome. FDR correction is further explained in the **Methods**.

Due to high missingness of anthropometric data at follow-up waves, BMI was only analyzed as an outcome at the baseline wave.
