## Supplemental Tables 5-8 for "Associations between prenatal caffeine exposure and child development: Longitudinal results from the Adolescent Brain Cognitive Development (ABCD) Study"

**Table S5.** Omnibus Results When Excluding Children With Non-Prevalent Substance Use

| **Outcome (n = 8969)** | $\chi^2$ | ***P* value** | **FDR-corrected P value** |
|---|---|---|---|
| Psychotic-like experiences | 5.82 | 0.12 | 0.32 |
| Internalizing per CBCL | 2.9 | 0.41 | 0.63 |
| Externalizing per CBCL | 9.15 | **0.03** | 0.12 |
| Attention per CBCL | 1.88 | 0.6 | 0.69 |
| Thought per CBCL | 0.36 | 0.95 | 0.95 |
| Social per CBCL | 2.55 | 0.47 | 0.63 |
| Body mass index | 12.1 | **7.04E-03** | 0.06 |
| Total sleep problems | 5.1 | 0.17 | 0.34 |

***Table S5 Note.*** Excluded children who reported using marijuana (n=72), using substances other than alcohol and tobacco (i.e., bath salts, n=2; "sniff" to "get high," n=9; inhalants, n=14; amphetamines not as prescribed, n=9; tranquilizers/anxiolytics/sedatives not as prescribed, n=11; pain medication not as prescribed, n=4; cough medication to "get high," n=8), or having a full drink of alcohol (n=123) or more than a puff of tobacco (n=36), or screened positive for substances based on hair toxicology (methamphetamines, n=11; THC, n=95; EtG, n=37; cotinine, n=9; Adderall, n=82). A total of 409 individuals were excluded for these analyses (Numbers do not sum as some children were positive across multiple indices). PLEs = Psychotic-Like Experiences. CBCL = Child Behavior Checklist. Due to high missingness of anthropometric data at follow-up waves, BMI was only analyzed as an outcome at the baseline wave.

**Table S6.** Omnibus Results When Excluding Children Prenatally Exposed to Illicit Substances Other Than Marijuana

| **Outcome (n = 8969)** | $\chi^2$ | ***P* value** | **FDR-corrected P value** |
|---|---|---|---|
| Psychotic-like experiences | 7.1 | 0.07 | 0.19 |
| Internalizing per CBCL | 3.07 | 0.38 | 0.51 |
| Externalizing per CBCL | 10.02 | **0.02** | 0.08 |
| Attention per CBCL | 3.04 | 0.39 | 0.51 |
| Thought per CBCL | 0.17 | 0.98 | 0.98 |
| Social per CBCL | 2.63 | 0.45 | 0.51 |
| Body mass index | 12.31 | **6.4E-03** | 0.05 |
| Total sleep problems | 6.13 | 0.11 | 0.22 |

***Table S6 Note.*** Children who were exposed to other illicit substances prenatally (i.e., cocaine or crack, n=51; heroin or morphine, n=19; oxycontin, n=25; other, n=33). A total of 102 individuals were excluded (numbers do not sum as some mothers were positive across multiple indices). PLEs

= Psychotic-Like Experiences. CBCL = Child Behavior Checklist. Due to high missingness of anthropometric data at follow-up waves, BMI was only analyzed as an outcome at the baseline wave.

**Table S7.** Omnibus Results When Excluding Children Born at Extreme Prematurity

| **Outcome (n = 8969)** | $\chi^2$ | ***P* value** | **FDR-corrected P value** |
|---|---|---|---|
| Psychotic-like experiences | 7.38 | 0.06 | 0.16 |
| Internalizing per CBCL | 2.76 | 0.43 | 0.5 |
| Externalizing per CBCL | 10.38 | **0.02** | 0.08 |
| Attention per CBCL | 2.71 | 0.44 | 0.5 |
| Thought per CBCL | 0.2 | 0.98 | 0.98 |
| Social per CBCL | 2.86 | 0.41 | 0.5 |
| Body mass index | 12.48 | **5.9E-03** | **0.047** |
| Total sleep problems | 5.27 | 0.15 | 0.3 |

***Table S7 Notes.*** Children born more than 8 weeks premature (i.e., less than 32 weeks' gestation; n=149) were excluded. PLEs = Psychotic-Like Experiences. CBCL = Child Behavior Checklist. Due to high missingness of anthropometric data at follow-up waves, BMI was only analyzed as an outcome at the baseline wave.

**Table S8.** Omnibus Results When Excluding Children Who Had a Non-Biological Mother Report as the Parent/Caregiver Respondent

| **Outcome (n = 8969)** | $\chi^2$ | ***P* value** | **FDR-corrected P value** |
|---|---|---|---|
| Psychotic-like experiences | 5.91 | 0.12 | 0.32 |
| Internalizing per CBCL | 3.78 | 0.29 | 0.46 |
| Externalizing per CBCL | 7.09 | 0.07 | 0.28 |
| Attention per CBCL | 1.32 | 0.73 | 0.77 |
| Thought per CBCL | 1.12 | 0.77 | 0.77 |
| Social per CBCL | 3.06 | 0.38 | 0.51 |
| Body mass index | 10.2 | **0.02** | 0.16 |
| Total sleep problems | 4.00 | 0.26 | 0.46 |

***Table S8 Notes.*** Children whose parent/caregiver respondent was not their biological mother were excluded (n=1,617). PLEs = Psychotic-Like Experiences. CBCL = Child Behavior Checklist. Due to high missingness of anthropometric data at follow-up waves, BMI was only analyzed as an outcome at the baseline wave.
